# The Role of Testing Availability on Intentions to Isolate during the COVID-19 Pandemic: A Randomized Trial

**DOI:** 10.1101/2021.03.22.21251380

**Authors:** Justin C. Zhang, Katherine L. Christensen, Richard K. Leuchter, Sitaram Vangala, Maria Han, Daniel M. Croymans

**Affiliations:** David Geffen School of Medicine at UCLA; Anderson School of Management at UCLA; UCLA Health Department of Medicine

**Keywords:** COVID-19, SARS-CoV-2, cross-sectional survey, testing, risky behavior, gender, health status, political affiliation

## Abstract

**Background:** Little information exists on how COVID-19 testing availability influences intentions to engage in risky behavior. Understanding the behavioral effects of testing availability may provide insight into the role of adequate testing on controlling viral transmission.

**Objective:** To evaluate the impact of testing availability on behavioral intention to self-isolate in a simulated scenario with participants who have been clinically diagnosed with COVID-19.

**Methods:** A total of 1400 participants were recruited from Amazon Mechanical Turk (MTurk) through a national, online, opt-in survey. Participants were randomized to one of three hypothetical scenarios. Each scenario asked participants to imagine having symptoms consistent with COVID-19 along with a clinical diagnosis from their physician. However, scenarios differed in their testing result: testing unavailable, positive test, or negative test. The primary outcome was intention to engage in high-risk COVID-19 behaviors, measured using an 11-item mean score (range 1-7) that was pre-registered prior to data collection. The randomized survey was conducted between July 23^rd^ to July 29^th^, 2020.

**Results:** Out of 1194 respondents (41.6% male, 58.4% female) with a median age of 38.5 years, participants who had no testing available in their clinical scenario showed significantly greater intentions to engage in behavior facilitating COVID-19 transmission compared to those who received a positive confirmatory test result scenario (difference (SE): 0.14 (0.06), *P*=0.016), equating to an 11.1% increase in mean score risky behavior intentions. Intention to engage in behaviors that can spread COVID-19 were also positively associated with male gender, poor health status, and Republican party affiliation.

**Conclusion:** Testing availability appears to play an independent role in influencing behaviors facilitating COVID-19 transmission. Such findings shed light on the possible negative externalities of testing unavailability.

**Trial Registration:** *Effect of Availability of COVID-19 Testing on Choice to Isolate and Socially Distance*, NCT04459520, https://clinicaltrials.gov/ct2/show/NCT04459520

## Introduction/Background

In the first six months of the coronavirus disease-2019 (COVID-19) pandemic, the U.S. has reported more cases of COVID-19 than any other country in the world [1]. One of the factors contributing to the spread of the SARS-Cov-2 virus has been inadequate self-isolation fueled by widespread testing unavailability affecting 40 of the 50 U.S. states [2]. Control of the virus has been further complicated by a polarized political landscape and longstanding sociodemographic disparities that limit access to healthcare [3,4].

While previous studies conducted during the COVID-19 pandemic as well as other infectious outbreaks have identified a number of factors associated with self-isolation behavior (e.g., demographics, vulnerability, political beliefs, fear), few studies have explored how COVID-19 confirmatory testing availability directly influences intentions to engage in behaviors which facilitate viral transmission [5-7]. Insight into this relationship would inform the public health response to future outbreaks. To this end, we conducted a web-based survey with randomized, hypothetical vignettes evaluating the impact of different testing scenarios on risky behavioral intentions in those presumed to have COVID-19.

## Methods

### Study Participants

Study participants were recruited from Amazon Mechanical Turk (MTurk), an online survey platform found to yield results comparable to that of traditional sampling means [8]. Participants were eligible if they were U.S. residents (defined by zip code), over 18 years of age, and passed all attention checks. Surveys were completed in Qualtrics. This randomized trial was exempted by the UCLA Institutional Review Board.

### Design

Eligible participants were invited to complete a 5-question, pre-test survey on Amazon Mechanical Turk, assessing Theory of Planned Behavior (TPB) construct response **(Supplemental eFigure 1)** and were compensated $ 0.10 for their time [9]. These questions assessed: individual attitudes, subjective norms, and perceived behavioral control regarding COVID-19 self-isolation and protective behaviors. An additional four questions were included for attention check purposes **(Supplemental eFigure 1)**. Participants who successfully completed the pre-test survey were invited to complete the full survey. To contact prior participants, we used the behavioral research tool, TurkPrime [8].

The survey instructed participants to imagine they were in one of three hypothetical scenarios. Each scenario began with the participant experiencing symptoms commonly associated with COVID-19 (fever and cough) and a physician clinically diagnosing them with COVID-19 and advising the participant to self-isolate. Each scenario differed in testing result: in arm 1 COVID-19 confirmatory testing was not available, in arm 2 the participant received a positive confirmatory test for the COVID-19 virus, and in arm 3 the participant received a negative confirmatory test for the COVID-19 virus.

After being presented with their respective scenarios, participants were then asked about their likelihood to engage in a number of behaviors over the following two weeks. These behaviors were primarily selected from those that the CDC identified as increasing the risk for contracting COVID-19 (hereinafter referred to as “risky behaviors”) [10,11]. Demographic questions and other risky behaviors of interest were included *a priori* (**Supplemental eFigure 2**) [12-16]. All main survey participants were compensated $ 0.60 for their time following survey completion.

Two pilot studies were run prior to launching the main survey to estimate power and effect size, as well as validate the internal consistency of the subscales **(Supplement eTable 1)**. The first pilot study results were used to develop an aggregate, 11-item risky behavior score, along with personal decisions and social expectations subscores. Three additional items (voting, protesting/counter-protesting, public transportation) were evaluated as individual scores. The second pilot was used to estimate an effect size (Cohen’s d) for comparing the testing not available condition with the testing positive condition in terms of the 11-item total score. The sample size of 1,194 (398 per condition) was chosen so as to provide 80% power to detect the estimated effect size of 0.23, assuming a two-sample t-test and a two-sided significance level of 0.017 (3-fold Bonferroni correction for pairwise comparison of study arms). Survey responses for the main study were collected between July 23^rd^, 2020 to July 29^th^, 2020.

Participants were excluded from the main study analysis if they were unable to complete the English language consent form, did not list a proper U.S. zip code, failed any one of the attention check questions in either the pre-test or the main survey, or completed the main survey in under 120 seconds—a threshold determined by the study team after pre-testing 15 college-educated individuals (**Supplemental eFigure 3**).

### Statistical Analysis

Survey responses were summarized for the full sample and stratified by testing scenario. Quantitative responses were summarized using means, standard deviations and quartiles, and categorical and ordinal responses were summarized using frequency distributions. Comparisons between scenarios were performed using one-way analysis of variance (ANOVA) for quantitative variables, Kruskal-Wallis tests for ordinal variables, and chi-squared or Fisher’s exact tests as appropriate for categorical variables.

The primary outcome was the 11-item mean score, ranging from 1 (minimal intention to engage in high-risk behavior) to 7 (maximal intention). Secondary outcomes were the personal decisions and social expectations subscales (each also ranging from 1 to 7), as well as the likelihood of voting, protesting/political gathering, and public transportation 1-item questions. Each outcome was analyzed using a pair of linear regression models. A pre-registered model included as covariates age, sex, race/ethnicity, political affiliation, education level, location, and type of residence. A post-hoc model included as additional covariates self-rated health status, the Consumer Finance Protection Bureau’s financial well-being score (financial well-being score), composite TPB score, region (metro adjacent v. not), and household risk. The financial well-being score is a 5-item score utilized by the U.S. government’s Consumer Finance Bureau to evaluate financial well-being [17]. Construct questions evaluating attitudes, subjective norms, and perceived behavioral control were adapted from a number of survey studies since no validated questionnaires specific to COVID-19 were available during the survey development period [9,18-19]. Because construct covariates had excellent internal consistency (a=0.83), they were aggregated into a single composite we refer to as the composite TPB score. For the pre-registered analysis of the primary outcome, pairwise comparisons of the 3 scenarios were performed using an 0.017 significance level (3-fold Bonferroni correction for an overall alpha of 0.05). All other analyses applied an 0.05 significance level. All analyses were performed using R v. 3.6.2 (http://www.r-project.org).

## Results

### Participants

Out of 1400 participants who completed the questionnaire, 1194 (85.3%) met all inclusion criteria and were included in the analysis. Basic demographic information presented in **Table 1** show that age, gender, race, and political affiliation were similar to national averages [20]. Participants were on average more educated than the national average. Arm 1 (testing unavailable group) contained 401 participants, arm 2 (positive confirmatory test group) contained 390 participants, and arm 3 (negative confirmatory test group) contained 403 participants.

**Table 1:** Participant Demographics.

| Participant Demographics |  |  |  |  |  |
| --- | --- | --- | --- | --- | --- |
|  |  | All Respondents (N=1194) | Arm 1 - Testing Unavailable (N=401) | Arm 2 - Positive Test (N=390) | Arm 3 – Negative Test (N=403) |
| Gender |  |  |  |  |  |
|  | Male | 497 (41.6%) | 159 (39.7%) | 169 (43.3%) | 169 (41.9%) |
|  | Female | 682 (57.1%) | 237 (59.1%) | 216 (55.4%) | 229 (56.8%) |
|  | Prefer to self-describe | 10 (0.8%) | 3 (0.7%) | 4 (1.0%) | 3 (0.7%) |
|  | Prefer not to say | 5 (0.4%) | 2 (0.5%) | 1 (0.3%) | 2 (0.5%) |
| Hispanic Ethnicity |  |  |  |  |  |
|  | Yes | 110 (9.2%) | 38 (9.5%) | 39 (10.0%) | 33 (8.2%) |
|  | No | 1069 (89.5%) | 358 (89.3%) | 346 (88.7%) | 365 (90.6%) |
|  | Prefer not to say | 15 (1.3%) | 5 (1.2%) | 5 (1.3%) | 5 (1.2%) |
| Race |  |  |  |  |  |
|  | White | 911 (79.0%) | 295 (76.6%) | 303 (80.2%) | 313 (80.3%) |
|  | Black or African American | 91 (7.9%) | 37 (9.6%) | 27 (7.1%) | 27 (6.9%) |
|  | Asian | 105 (9.1%) | 39 (10.1%) | 33 (8.7%) | 33 (8.5%) |
|  | Native Hawaiian or Pacific Islander | 2 (0.2%) | 1 (0.3%) | 1 (0.3%) | 0 |
|  | American Indian or Alaskan | 10 (0.9%) | 2 (0.5%) | 2 (0.5%) | 6 (1.5%) |
|  | Native |  |  |  |  |
|  | Some other race, ethnicity, or origin | 20 (1.7%) | 6 (1.6%) | 8 (2.1%) | 6 (1.5%) |
|  | Prefer not to say | 14 (1.2%) | 5 (1.3%) | 4 (1.1%) | 5 (1.3%) |
|  | Missing | 41 | 16 | 12 | 13 |
| Political Affiliation |  |  |  |  |  |
|  | Republican | 277 (23.2%) | 86 (21.4%) | 78 (20.0%) | 113 (28.0%) |
|  | Democrat | 528 (44.2%) | 177 (44.1%) | 175 (44.9%) | 176 (43.7%) |
|  | Independent | 389 (32.6%) | 138 (34.4%) | 137 (35.1%) | 114 (28.3%) |
| Level of Education |  |  |  |  |  |
|  | 8th grade or less | 1 (0.1%) | 1 (0.2%) | 0 | 0 |
|  | Some high school, but did not graduate | 8 (0.7%) | 4 (1.0%) | 3 (0.8%) | 1 (0.2%) |
|  | High school graduate or GED | 131 (11.0%) | 49 (12.2%) | 42 (10.8%) | 40 (9.9%) |
|  | Some college or 2-year degree | 355 (29.7%) | 109 (27.2%) | 124 (31.8%) | 122 (30.3%) |
|  | 4-year college degree | 451 (37.8%) | 155 (38.7%) | 130 (33.3%) | 166 (41.2%) |
|  | More than 4-year college degree | 248 (20.8%) | 83 (20.7%) | 91 (23.3%) | 74 (18.4%) |
| Type of Residence |  |  |  |  |  |
|  | House/condo/townhouse | 910 (76.2%) | 313 (78.1%) | 298 (76.4%) | 299 (74.2%) |
|  | Apartment | 271 (22.7%) | 83 (20.7%) | 88 (22.6%) | 100 (24.8%) |
|  | Dormitory | 1 (0.1%) | 0 | 0 | 1 (0.2%) |
|  | Assisted living facility | 1 (0.1%) | 1 (0.2%) | 0 | 0 |
|  | Other | 11 (0.9%) | 4 (1.0%) | 4 (1.0%) | 3 (0.7%) |
| Shared Living Space |  |  |  |  |  |
|  | I live by myself | 186 (15.6%) | 66 (16.5%) | 62 (15.9%) | 58 (14.4%) |
|  | 2 people | 406 (34.0%) | 127 (31.7%) | 150 (38.5%) | 129 (32.0%) |
|  | 3 people | 243 (20.4%) | 84 (20.9%) | 75 (19.2%) | 84 (20.8%) |
|  | 4 people | 212 (17.8%) | 74 (18.5%) | 57 (14.6%) | 81 (20.1%) |
|  | 5 people | 97 (8.1%) | 28 (7.0%) | 32 (8.2%) | 37 (9.2%) |
|  | 6 or more people | 50 (4.2%) | 22 (5.5%) | 14 (3.6%) | 14 (3.5%) |
| Overall Health |  |  |  |  |  |
|  | Excellent | 183 (15.3%) | 73 (18.2%) | 48 (12.3%) | 62 (15.4%) |
|  | Very good | 491 (41.1%) | 166 (41.4%) | 173 (44.4%) | 152 (37.7%) |
|  | Good | 382 (32.0%) | 111 (27.7%) | 126 (32.3%) | 145 (36.0%) |
|  | Fair | 119 (10.0%) | 46 (11.5%) | 36 (9.2%) | 37 (9.2%) |
|  | Poor | 19 (1.6%) | 5 (1.2%) | 7 (1.8%) | 7 (1.7%) |
| Household Member under 18 |  |  |  |  |  |
|  | Yes | 419 (35.1%) | 147 (36.7%) | 117 (30.0%) | 155 (38.5%) |
|  | No | 775 (64.9%) | 254 (63.3%) | 273 (70.0%) | 248 (61.5%) |
| Household Vulnerability |  |  |  |  |  |
|  | Yes | 353 (29.6%) | 128 (31.9%) | 112 (28.7%) | 113 (28.0%) |
|  | No | 829 (69.4%) | 268 (66.8%) | 274 (70.3%) | 287 (71.2%) |
|  | Do not know | 7 (0.6%) | 3 (0.7%) | 3 (0.8%) | 1 (0.2%) |
|  | Prefer not to say | 5 (0.4%) | 2 (0.5%) | 1 (0.3%) | 2 (0.5%) |

### Relationship of COVID-19 Test Result to Behavioral Intentions: Primary Outcome

The analysis of the primary outcome (**Table 2**) indicated that testing unavailability resulted in a significantly greater self-reported intention to engage in risky behavior compared to those with a positive test (arm 2) (difference (SE): 0.14 (0.06), *P*=0.016). This difference corresponds to an 11% relative increase in risky behavior intentions based on mean intention scores (**Figure 1**). Participants with negative tests demonstrated the greatest intention to engage in risky behavior compared to those without available testing (difference (SE): 0.35 (0.06), *P*<0.001) and positive tests (difference (SE): 0.49 (0.06), *P*<0.001), respectively. Relative to positive test group’s mean score, those who received a negative test were 39% more likely to engage in risky behavior. Similar significant differences were noted when comparing the personal decisions and the social expectations subscores.

**Table 2:** **Linear Regression Model Evaluating Associations between Behavioral Intentions and Pre-Specified Covariates**

| Effect | Total Score ($R^2 = 0.11$) | | | Personal Score ($R^2 = 0.10$) | | | Social Score ($R^2 = 0.10$) | | |
| --- | --- | --- | --- | --- | --- | --- | --- | --- | --- |
|  | Estimate | SE | P | Estimate | SE | P | Estimate | SE | P |
| Scenario 2 v. 1 | -0.14 | 0.06 | **0.016** | -0.12 | 0.06 | **0.045** | -0.18 | 0.07 | **0.011** |
| Scenario 3 v. 1 | 0.35 | 0.06 | **<0.001** | 0.31 | 0.06 | **<0.001** | 0.42 | 0.07 | **<0.001** |
| Scenario 3 v. 2 | 0.49 | 0.06 | **<0.001** | 0.42 | 0.06 | **<0.001** | 0.60 | 0.07 | **<0.001** |
| Age (+1y) | 0.00 | 0.00 | **0.036** | 0.00 | 0.00 | 0.177 | -0.01 | 0.00 | **0.006** |
| Gender (Ref = Male) |  |  | **0.024** |  |  | **0.011** |  |  | 0.137 |
| Female | -0.13 | 0.05 | **0.009** | -0.15 | 0.05 | **0.002** | -0.09 | 0.06 | 0.147 |
| Prefer to self-describe | -0.34 | 0.29 | 0.228 | -0.25 | 0.28 | 0.371 | -0.50 | 0.35 | 0.157 |
| Prefer not to say | -0.65 | 0.42 | 0.124 | -0.59 | 0.42 | 0.162 | -0.75 | 0.52 | 0.148 |
| Race/Ethnicity (Ref = White) |  |  | 0.854 |  |  | 0.316 |  |  | 0.897 |
| Hispanic | -0.06 | 0.09 | 0.458 | -0.09 | 0.09 | 0.289 | -0.02 | 0.11 | 0.875 |
| Black or African American | 0.04 | 0.09 | 0.633 | 0.10 | 0.09 | 0.266 | -0.06 | 0.12 | 0.613 |
| Asian | -0.04 | 0.09 | 0.607 | -0.12 | 0.09 | 0.150 | 0.09 | 0.11 | 0.376 |
| Native Hawaiian or Pacific Islander | 0.42 | 0.80 | 0.596 | 0.94 | 0.79 | 0.239 | -0.47 | 0.99 | 0.632 |
| American Indian or Alaskan Native | -0.37 | 0.33 | 0.261 | -0.34 | 0.33 | 0.291 | -0.41 | 0.41 | 0.313 |
| Some other race, ethnicity, or origin | -0.06 | 0.30 | 0.837 | 0.00 | 0.30 | 0.995 | -0.17 | 0.38 | 0.654 |
| Prefer not to say | 0.21 | 0.27 | 0.447 | 0.23 | 0.27 | 0.385 | 0.16 | 0.34 | 0.641 |
| Political Affiliation (Ref = Republican) |  |  | **<0.001** |  |  | **<0.001** |  |  | **<0.001** |
| Democrat | -0.36 | 0.06 | **<0.001** | -0.32 | 0.06 | **<0.001** | -0.42 | 0.08 | **<0.001** |
| Independent | -0.13 | 0.07 | **0.041** | -0.15 | 0.07 | **0.021** | -0.10 | 0.08 | 0.197 |
| Education (Ref = 8th grade or less) |  |  | 0.587 |  |  | 0.445 |  |  | 0.861 |
| Some high school, but did not graduate | 0.07 | 0.85 | 0.935 | -0.20 | 0.84 | 0.816 | 0.53 | 1.05 | 0.611 |
| High school graduate or GED | 0.07 | 0.80 | 0.929 | -0.17 | 0.80 | 0.835 | 0.49 | 0.99 | 0.623 |
| Some college or 2-year degree | 0.16 | 0.80 | 0.838 | -0.07 | 0.79 | 0.930 | 0.57 | 0.99 | 0.562 |
| 4-year college degree | 0.14 | 0.80 | 0.865 | -0.11 | 0.79 | 0.891 | 0.56 | 0.99 | 0.568 |
| More than 4-year college degree | 0.23 | 0.80 | 0.775 | 0.00 | 0.80 | 0.999 | 0.62 | 0.99 | 0.528 |
| Residence (Ref = House/condo/townhouse) |  |  | 0.807 |  |  | 0.600 |  |  | 0.931 |
| Apartment | -0.07 | 0.06 | 0.228 | -0.09 | 0.06 | 0.111 | -0.03 | 0.07 | 0.661 |
| Dormitory | -0.24 | 0.80 | 0.770 | 0.10 | 0.80 | 0.901 | -0.82 | 1.00 | 0.410 |
| Assisted Living Facility | -0.10 | 0.80 | 0.898 | -0.16 | 0.80 | 0.843 | -0.01 | 0.99 | 0.995 |
| Other | -0.08 | 0.26 | 0.747 | -0.12 | 0.26 | 0.641 | -0.02 | 0.32 | 0.952 |

Pre-registered linear regression analysis based on the pre-specified covariates. Estimate refers to mean estimate difference compared to the reference group, and SE refers to Standard Error. Statistically significant differences were shown through bolded p-values.

**Figure 1:**
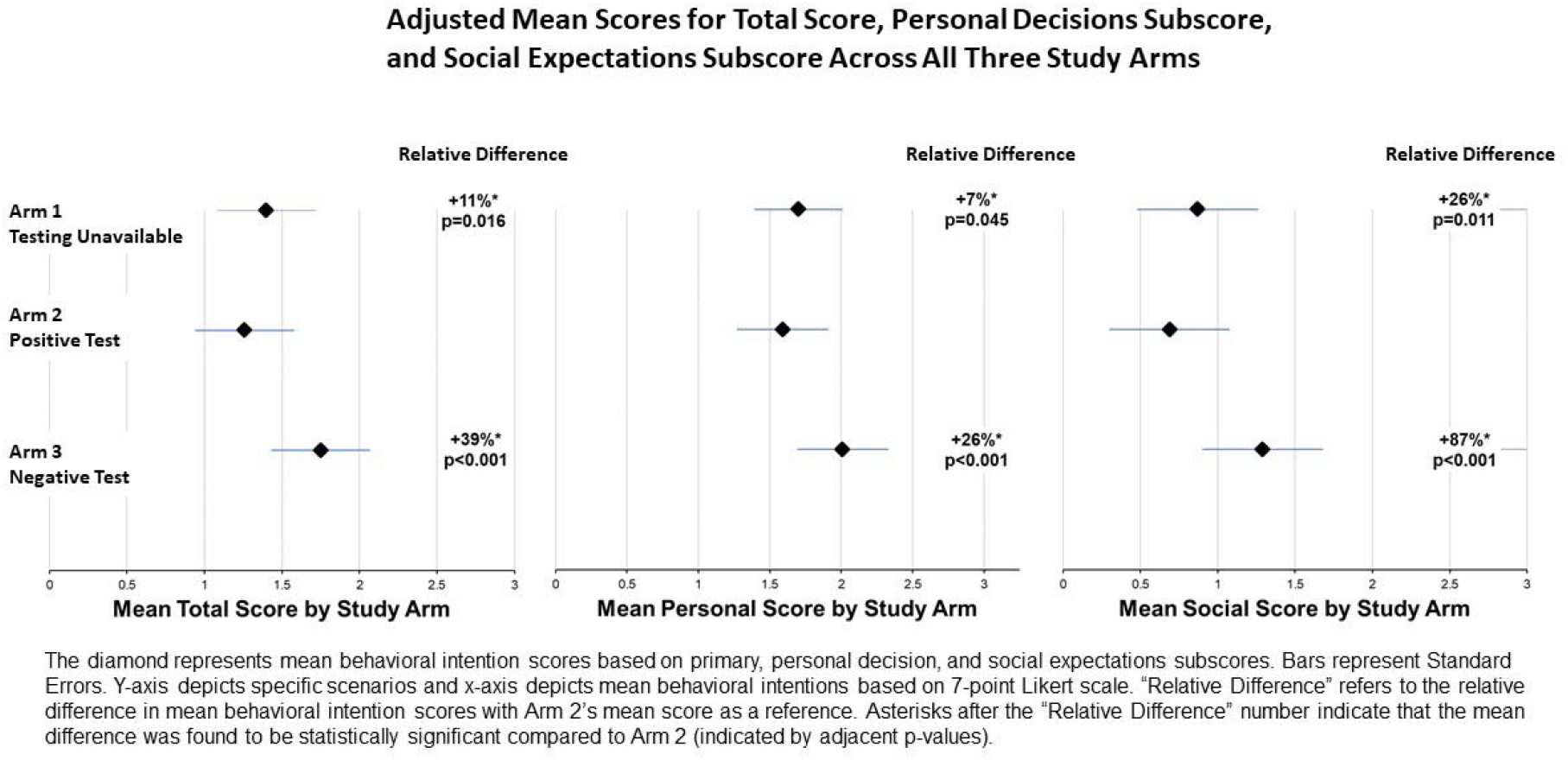
**Adjusted Mean Scores for Total Score, Personal Decisions Subscore, and Social Expectations Subscore Across All Three Study Arms**

### Linear Regression Modelling

The pre-registered model (**Table 2**) included several pre-specified covariates: gender, race, ethnicity, political affiliation, level of education, and type of residence. This model explained only 11% of the variance in the primary outcome. As a result, a post-hoc model **(Table 3)** incorporating several additional covariates (health status, financial well-being, composite TPB score, metro code, and household risk) was developed and found to explain 35% of the primary outcome variance. Exploratory analyses suggest that the R^2^ of the pre-specified model was largely driven by scenario differences and political affiliation, while the R^2^ for the post-hoc model was largely driven by scenario differences, political affiliation, and composite TPB scores.

**Table 3:** **Post-Hoc Linear Regression Model Evaluating Associations between Behavioral Intentions (Total Score, Subscores, Voting, and Protest) and Additional Covariates**

| Effect | Total Score ($R^2 = 0.35$) | | | Personal Score ($R^2 = 0.32$) | | | Social Score ($R^2 = 0.29$) | | | Voting ($R^2 = 0.16$) | | | Protest ($R^2 = 0.15$) | | |
| --- | --- | --- | --- | --- | --- | --- | --- | --- | --- | --- | --- | --- | --- | --- | --- |
|  | Estimate | SE | P | Estimate | SE | P | Estimate | SE | P | Estimate | SE | P | Estimate | SE | P |
| Scenario 2 v. 1 | -0.17 | 0.05 | **<0.001** | -0.15 | 0.05 | **0.003** | -0.22 | 0.06 | **<0.001** | -0.33 | 0.12 | **0.006** | -0.11 | 0.04 | **0.012** |
| Scenario 3 v. 1 | 0.32 | 0.05 | **<0.001** | 0.28 | 0.05 | **<0.001** | 0.38 | 0.06 | **<0.001** | 0.39 | 0.12 | **0.001** | 0.11 | 0.04 | **0.011** |
| Scenario 3 v. 2 | 0.50 | 0.05 | **<0.001** | 0.44 | 0.05 | **<0.001** | 0.60 | 0.06 | **<0.001** | 0.73 | 0.12 | **<0.001** | 0.22 | 0.04 | **<0.001** |
| Age (+1y) | 0.00 | 0.00 | 0.792 | 0.00 | 0.00 | 0.596 | 0.00 | 0.00 | 0.199 | 0.01 | 0.00 | 0.091 | 0.00 | 0.00 | 0.408 |
| Gender (Ref = Male) |  |  | 0.130 |  |  | 0.105 |  |  | 0.213 |  |  | 0.534 |  |  | 0.788 |
| Female | -0.06 | 0.04 | 0.163 | -0.08 | 0.04 | **0.047** | -0.01 | 0.05 | 0.798 | -0.13 | 0.10 | 0.191 | 0.03 | 0.04 | 0.348 |
| Prefer to self-describe | -0.38 | 0.25 | 0.121 | -0.29 | 0.25 | 0.243 | -0.55 | 0.32 | 0.088 | -0.27 | 0.60 | 0.651 | 0.09 | 0.21 | 0.688 |
| Prefer not to say | -0.54 | 0.38 | 0.157 | -0.49 | 0.38 | 0.203 | -0.63 | 0.49 | 0.202 | -0.67 | 0.93 | 0.473 | -0.07 | 0.33 | 0.836 |
| Race/Ethnicity (Ref = White) |  |  | 0.725 |  |  | 0.176 |  |  | 0.798 |  |  | 0.957 |  |  | 0.798 |
| Hispanic | -0.07 | 0.07 | 0.346 | -0.10 | 0.08 | 0.208 | -0.03 | 0.10 | 0.773 | -0.20 | 0.18 | 0.281 | 0.04 | 0.07 | 0.546 |
| Black or African American | 0.05 | 0.08 | 0.554 | 0.11 | 0.08 | 0.192 | -0.05 | 0.10 | 0.602 | -0.05 | 0.19 | 0.783 | 0.01 | 0.07 | 0.919 |
| Asian | 0.00 | 0.07 | 0.963 | -0.07 | 0.07 | 0.322 | 0.14 | 0.10 | 0.147 | -0.15 | 0.18 | 0.397 | 0.02 | 0.06 | 0.716 |
| Native Hawaiian or Pacific Islander | 0.84 | 0.69 | 0.220 | 1.35 | 0.69 | 0.052 | -0.04 | 0.89 | 0.961 | -0.43 | 1.67 | 0.797 | 0.10 | 0.60 | 0.864 |
| American Indian or Alaskan Native | -0.34 | 0.28 | 0.226 | -0.34 | 0.29 | 0.238 | -0.35 | 0.37 | 0.335 | -0.28 | 0.69 | 0.688 | 0.06 | 0.25 | 0.804 |
| Some other race, ethnicity, or origin | 0.06 | 0.26 | 0.817 | 0.13 | 0.26 | 0.618 | -0.06 | 0.34 | 0.852 | 0.06 | 0.64 | 0.926 | 0.13 | 0.23 | 0.557 |
| Prefer not to say | 0.03 | 0.24 | 0.894 | 0.05 | 0.24 | 0.841 | 0.00 | 0.30 | 0.991 | 0.17 | 0.57 | 0.760 | 0.37 | 0.20 | 0.074 |
| Political Affiliation (Ref = Republican) |  |  | 0.082 |  |  | 0.364 |  |  | **0.014** |  |  | 0.437 |  |  | 0.135 |
| Democrat | -0.10 | 0.06 | 0.089 | -0.07 | 0.06 | 0.199 | -0.14 | 0.07 | 0.061 | -0.10 | 0.14 | 0.452 | 0.00 | 0.05 | 0.958 |
| Independent | 0.00 | 0.06 | 0.960 | -0.02 | 0.06 | 0.737 | 0.04 | 0.07 | 0.572 | -0.18 | 0.14 | 0.199 | 0.08 | 0.05 | 0.113 |
| Education (Ref = 8th grade or less) |  |  | 0.234 |  |  | 0.233 |  |  | 0.423 |  |  | 0.208 |  |  | 0.848 |
| Some high school, but did not graduate | 0.69 | 0.73 | 0.348 | 0.39 | 0.74 | 0.602 | 1.22 | 0.95 | 0.199 | 1.21 | 1.78 | 0.497 | 0.49 | 0.64 | 0.445 |
| High school graduate or GED | 0.53 | 0.69 | 0.445 | 0.26 | 0.70 | 0.705 | 0.99 | 0.90 | 0.268 | 1.03 | 1.68 | 0.542 | 0.45 | 0.60 | 0.458 |
| Some college or 2-year degree | 0.66 | 0.69 | 0.339 | 0.39 | 0.70 | 0.579 | 1.14 | 0.90 | 0.202 | 1.27 | 1.68 | 0.449 | 0.40 | 0.60 | 0.504 |
| 4-year college degree | 0.61 | 0.69 | 0.376 | 0.33 | 0.70 | 0.639 | 1.11 | 0.90 | 0.214 | 1.33 | 1.68 | 0.428 | 0.40 | 0.60 | 0.503 |
| More than 4-year college degree | 0.70 | 0.69 | 0.312 | 0.44 | 0.70 | 0.533 | 1.17 | 0.90 | 0.193 | 1.50 | 1.68 | 0.372 | 0.45 | 0.60 | 0.457 |
| Health Status (Ref = "Excellent") |  |  | **0.037** |  |  | **0.028** |  |  | **0.033** |  |  | **0.026** |  |  | 0.947 |
| Very good | -0.13 | 0.06 | **0.037** | -0.06 | 0.06 | 0.330 | -0.25 | 0.08 | **0.002** | -0.46 | 0.15 | **0.002** | 0.03 | 0.05 | 0.613 |
| Good | -0.15 | 0.07 | **0.021** | -0.11 | 0.07 | 0.104 | -0.23 | 0.08 | **0.007** | -0.37 | 0.16 | **0.018** | 0.03 | 0.06 | 0.649 |
| Fair | -0.09 | 0.09 | 0.307 | -0.03 | 0.09 | 0.739 | -0.20 | 0.11 | 0.086 | -0.44 | 0.21 | **0.040** | -0.01 | 0.08 | 0.897 |
| Poor | -0.45 | 0.17 | **0.008** | -0.52 | 0.17 | **0.003** | -0.33 | 0.22 | 0.135 | -0.81 | 0.42 | 0.051 | -0.04 | 0.15 | 0.782 |
| CFPB Score (+1) | 0.00 | 0.00 | 0.635 | 0.00 | 0.00 | 0.431 | 0.00 | 0.00 | 0.947 | 0.00 | 0.00 | 0.555 | 0.00 | 0.00 | 0.342 |
| Construct Score (+1) | -0.12 | 0.01 | **<0.001** | -0.11 | 0.01 | **<0.001** | -0.12 | 0.01 | **<0.001** | -0.16 | 0.01 | **<0.001** | -0.06 | 0.01 | **<0.001** |
| Description (Ref = Metro) |  |  | **0.025** |  |  | 0.082 |  |  | **0.022** |  |  | 0.562 |  |  | 0.220 |
| Nonmetro - metro adjacent | 0.21 | 0.08 | **0.007** | 0.17 | 0.08 | **0.029** | 0.28 | 0.10 | **0.006** | 0.20 | 0.19 | 0.309 | 0.11 | 0.07 | 0.109 |
| Nonmetro - not metro adjacent | 0.06 | 0.10 | 0.534 | 0.07 | 0.10 | 0.516 | 0.06 | 0.13 | 0.661 | 0.11 | 0.25 | 0.670 | 0.07 | 0.09 | 0.419 |
| Household Risk (Ref = Yes) |  |  | 0.110 |  |  | 0.113 |  |  | 0.190 |  |  | 0.476 |  |  | 0.884 |
| No | 0.09 | 0.05 | 0.054 | 0.07 | 0.05 | 0.132 | 0.13 | 0.06 | **0.040** | -0.14 | 0.12 | 0.226 | 0.00 | 0.04 | 0.927 |
| Do not know | 0.36 | 0.27 | 0.179 | 0.48 | 0.27 | 0.078 | 0.16 | 0.35 | 0.646 | 0.56 | 0.65 | 0.393 | -0.09 | 0.23 | 0.686 |
| Prefer not to say | 0.43 | 0.34 | 0.210 | 0.43 | 0.34 | 0.206 | 0.42 | 0.44 | 0.345 | -0.16 | 0.83 | 0.842 | -0.21 | 0.30 | 0.479 |
| Residence (Ref = House/condo/townhouse) |  |  | 0.200 |  |  | 0.097 |  |  | 0.684 |  |  | 0.182 |  |  | 0.589 |
| Apartment/assisted living facility/dormito | -0.09 | 0.05 | 0.078 | -0.11 | 0.05 | **0.033** | -0.06 | 0.07 | 0.393 | -0.20 | 0.12 | 0.108 | -0.04 | 0.04 | 0.411 |
| Other | -0.09 | 0.22 | 0.674 | -0.11 | 0.23 | 0.619 | -0.06 | 0.29 | 0.827 | -0.54 | 0.54 | 0.317 | -0.13 | 0.19 | 0.506 |

Post-hoc linear regression analysis, which includes additional covariates (health status, zip code density, CFPB, and composite TPB score) compared to the pre-registered model. Estimate refers to mean estimate difference compared to the reference group, and SE refers to Standard Error. Statistically significant differences were shown through bolded p-values.

### Gender

Our pre-specified model found that gender identification was significantly associated with risky behavior intentions across the primary outcome and the two subscores. When compared to those who identify as men, those identifying as women reported significantly decreased risky behavior intentions (difference (SE): −0.13 (0.05), *P*=0.009) as measured by the primary outcome (**Table 2**). However, in the post-hoc model that included a control for pre-existing attitudes about the disease (the composite TPB score), this gender difference in behavior intentions was non-significant (difference (SE): −0.06 (0.04), *P*=0.163) (**Table 3**).

### Health Status

After controlling for other variables in the post-hoc model, self-identified health status was found to be significantly associated with behavioral intentions. Compared to those who reported “excellent” health status, those who reported a “poor” health status expressed the lowest intentions to engage in risky behavior (difference (SE): −0.45 (0.17), *P*=0.008) (**Table 3**).

### Political Affiliation

In the pre-specified model, Republicans reported higher intentions to engage in risky behavior than either Democrats (difference (SE): 0.36 (0.06), *P*<0.001) or Independents (difference (SE): − 0.13 (0.07), *P*=0.041), respectively. In other words, Republicans showed a 27% relative increase in mean intention score to engage in risky behavior compared to Democrats. Republicans were also less likely than Democrats and Independents to agree with the statement that “COVID-19 could have severe consequences on other peoples’ lives”. In the post-hoc model that included a control for pre-existing attitudes about the severity of the disease, the effect of political identity on intention to self-isolate was no longer significant.

### Voting and Rally / Protest Intentions

Since in-person voting for the United States presidential election will occur this November, we assessed voting intentions and intention to participate in a large-scale political event (a rally or a protest). Participants who received a positive test result indicated a lower intention to vote in person than participants who had not received confirmatory testing (difference (SE): −0.30 (0.13), *P*=0.018). Participants who tested negative indicated higher intentions to participate in a rally or protest than both participants who tested positive (difference (SE): 0.21 (0.05), *P*<0.001) and participants who did not receive a test result (difference (SE): 0.13 (0.05), *P*=0.005).

Political affiliation also affected intention to vote in-person and intention to attend a rally or protest. Democrats reported decreased intentions to cast a ballot at a voting station (difference (SE): −0.43 (0.14), *P*=0.002) and attend a rally, protest or counter-protest (difference (SE): −0.13 (0.05), *P*=0.008) compared to Republican respondents. In other words, Democrats demonstrated a 33% and 12% relative decrease in mean intention scores to vote in-person and attend a protest/political rally respectively. Those identifying as politically independent indicated significantly decreased intentions to vote in-person (difference (SE): −0.34 (0.14), *P*=0.017) compared to their Republican counterparts, but were not significantly less likely to attend a rally or a protest.

### The Theory of Planned Behavior

The notable difference in R^2^ between the pre-registered (R^2^=0.11) and post-hoc (R^2^=0.36) linear regression models suggest that the composite TPB score plays a significant role in explaining the variance seen with the total score. When evaluated within the post-hoc model, the first subjective norms question, “*People with COVID-19 should self-isolate*” showed the largest negative association with risky behavioral intentions (difference (SE): −0.27 (0.05), *P*<0.001) (**Supplemental eTable 3**). Both attitudes questions demonstrated significant, yet smaller effect sizes (difference (SE): −0.12 (0.05), *P*=0.009) (difference (SE): −0.15 (0.03), *P*<0.001), and perceived behavioral control demonstrated a weaker, but nonetheless significant effect (difference (SE): −0.05 (0.02), *P*=0.022).

## Discussion

Controlling for key demographic variables, participants who were clinically diagnosed with COVID-19 but had no testing available to them exhibited an 11% relative increase in intention to engage in risky behavior compared to those with a positive confirmatory test. Additionally, clinically symptomatic participants who received a negative test reported higher intentions to engage in risky behavior than any other group. The present findings are relevant to both the outbreak and continued transmission of COVID-19 given that a majority of states do not have sufficient testing availability [2]. This study is the first to demonstrate how testing unavailability independently decreases intention to isolate in patients clinically diagnosed with COVID-19. While increasing testing availability alone will not fully eliminate viral transmission, recent evidence indicates that relatively small degrees of behavioral change (e.g., decreasing visits to non-essential businesses, wearing masks) may result in major decreases in viral transmission [21,22]. Thus, while the magnitude of behavior intention change reported in this study is small on an absolute level, our findings nonetheless suggest a clear role that testing availability could play in curbing viral transmission. These findings are particularly timely as the U.S. experiences a resurgence of cases [1].

In addition to the impact of testing unavailability, it is interesting to note the impact a negative test has on increasing risky behavior intentions. Despite a clinical diagnosis, those with a negative confirmatory test were significantly more likely to engage in behaviors facilitating viral transmission, likely because they believed that they could not transmit the virus to others. However, the larger magnitude of increased intention to engage in risky behavior among those with negative confirmatory tests relative to the other groups may be clinically relevant given reports of false negatives and testing uncertainty throughout the lifecycle of the virus [23,24].

The present results are also notable for the effect that political leanings have on the intention to self-isolate. Prior studies have found a correlation between mistrust of government-issued guidelines and partisan affiliation [25]. This study finds that Republicans are not only less likely to agree that “*COVID-19 could have severe consequences on other peoples’ lives*,”, but also are 27% more likely to engage in risky behaviors compared to Democrats based on mean intention scores. These findings indicate that decreased belief in the dangers of the disease may play a role in Republicans’ decision to self-isolate and may suggest a potential pathway by which partisan beliefs influence behavioral intentions.

The results presented here also suggest that political leanings affect both intentions to vote in-person and intentions to attend political events that involve crowds (e.g., rally, protest). Regardless of testing availability, Democrats with a presumed COVID-19 diagnosis indicated a remarkable 33% lower likelihood to engage in in-person voting based on their respective mean intention scores. One downstream implication of these exploratory results may be that communities burdened with high rates of COVID-19 infections during Election Day might have significantly lower Democratic than Republican on-site turnout.

We found that those with poorer health revealed greater intentions to self-isolate. While several studies have shown that increased risk perception is associated with adopting self-protective behaviors, none of them evaluated the effect on behaviors post-infection [26-28]. It may be possible that those with greater comorbidities are particularly sensitive to the negative impact of transmitting the infection to others within their community. If so, these findings suggest that healthy people may inadvertently view social isolation and protective behaviors as an individual choice as opposed to those in poor health who may view it instead as a social obligation.

The fact that the Theory of Planned Behavior explains a significant amount of variance within our models suggests that changing public perception regarding the severity of the virus, subjective norms, or the perceived self-efficacy to engage in protective behavior can serve as a strategy to decrease risky behavior.

Although not directly evaluated here, it may be interesting to consider the effect of waiting to receive a definitive result on intention to engage in risky behaviors. While we hypothesize that a delay in test results might lead patients to behave similarly to respondents assigned to arm 1, further studies are necessary to evaluate the effect of testing delay on self-isolation behavior.

### Limitations

There are several limitations to our study. First, survey respondents were recruited using MTurk, an online platform that—while as effective as traditional survey sampling methods— skews towards younger, more well-educated individuals [29]. Thus, our participant cohort cannot be interpreted as a nationally representative sample. Nonetheless, given the current pandemic and the need to minimize viral exposure for both researchers and participants, surveying study participants remotely using a well-established, online platform seemed the most reasonable and ethical means to conduct our study.

Study findings may also be limited by the hypothetical nature of the survey design, where we asked participants to imagine how they would react in such a scenario. However, survey scenarios were designed to be easily readable and were repeated on several pages in order to facilitate participant comprehension and immersion (**Supplement eFigure 4**). To further reduce the number of participants who did not take an appropriate amount of time to imagine the scenario, we restricted analysis to participants who spent at least 2 minutes on the survey and passed all attention checks.

External validity of this study is limited by the fact that the study only evaluated behavioral intentions. A large body of prior research has noted that behavioral intentions do not immediately translate to behavioral engagement but are rather attenuated or enhanced by other factors. Nevertheless, behavioral intentions are commonly acknowledged as one of the best predictors of behaviors themselves, so it is likely that differences in behavior between the different groups would still exist [30,31].

Lastly, given the dynamic nature of the global pandemic and the recent shifts in public opinion towards both the global pandemic and self-protective measures, public opinion on this subject will likely continue to shift over time [32]. Despite these limitations, these significant differences in behavioral intentions are novel findings providing evidence that increased testing capacity may ultimately translate into fewer infections and fewer deaths.

## Conclusion

Testing availability independently influences patients’ intentions to engage in COVID-19 risky behaviors, even when controlling for a clinical diagnosis of COVID-19. Such findings shed light on the possible negative externalities of testing unavailability and thus highlight how increased testing availability might directly translate to decreased infections and deaths.

## Data Availability

We have archived participant data from the online surveys. This data is available upon request.

## Acknowledgements

We would like to thank The Morrison Center for Marketing and Data Analytics and UCLA Health Department of Medicine for providing financial funding allowing for us to conduct this survey study.

## Author Contributions

Dr. Daniel M. Croymans, Dr. Richard K. Leuchter, Justin C. Zhang, Sitaram Vangala, Katherine L. Christensen, and Maria Han had full access to all of the data in the study and take responsibility for the integrity of the data and the accuracy of the data analysis.

*Concept and design:* All authors.

*Acquisition, analysis, or interpretation of data:* Justin C. Zhang, Daniel M. Croymans, Katherine L. Christensen, Richard K. Leuchter

*Drafting of the manuscript:* All authors.

*Critical revision of the manuscript for important intellectual content:* Justin C. Zhang, Daniel M. Croymans, Katherine L. Christensen, Richard K. Leuchter

*Statistical analysis:* Sitaram Vangala

*Obtained funding:* Katherine L. Christensen, Daniel M. Croymans

*Administrative, technical, or material support:* Sitaram Vangala

*Supervision:* Daniel M. Croymans

