## Supplementary material for "The Role of Testing Availability on Intentions to Isolate during the COVID-19 Pandemic: A Randomized Trial": Figure 1

Supplemental Document

**Supplemental eFigure 1:** Pre-test Survey (Qualtrics Printout)

COVID-19 Pre-Test

Start of Block

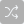

Q20 This is a nine question, one-page study. Thanks for your help!

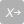

[attitude1] If I have active COVID-19, engaging in self-isolation behaviors will reduce the spread of the virus to others.^3^

- Strongly disagree (1)
- Disagree (2)
- Somewhat disagree (3)
- Neither agree nor disagree (4)
- Somewhat agree (5)
- Agree (6)
- Strongly agree (7)

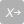

[attitude2] COVID-19 could have severe consequences on other peoples’ lives.^3^

- Strongly disagree (1)
- Disagree (2)
- Somewhat disagree (3)
- Neither agree nor disagree (4)
- Somewhat agree (5)
- Agree (6)
- Strongly agree (7)

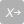

[subjnorm1] People with COVID-19 should self-isolate.^3^

- Strongly disagree (1)
- Disagree (2)
- Somewhat disagree (3)
- Neither agree nor disagree (4)
- Somewhat agree (5)
- Agree (6)
- Strongly agree (7)

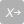

[subjnorm2] Most people who are important to me think that I should self-isolate if infected with COVID-19.^3^

- Strongly disagree (1)
- Disagree (2)
- Somewhat disagree (3)
- Neither agree nor disagree (4)
- Somewhat agree (5)
- Agree (6)
- Strongly agree (7)

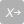

[control] If I have an active COVID-19 infection, I am confident that I could self-isolate.^3^

- Strongly disagree (1)
- Disagree (2)
- Somewhat disagree (3)
- Neither agree nor disagree (4)
- Somewhat agree (5)
- Agree (6)
- Strongly agree (7)

| 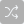 | 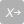 |
| --- | --- |

[crying] When someone is crying which of these is most likely how they feel?

- Sad1 (1)
- 2 (2)
- 3 (3)
- 4 (4)
- 5 (5)
- 6 (6)
- Happy7 (7)

| 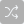 | 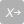 |
| --- | --- |

[smile] When someone is smiling which of these is most likely how they feel?

- Sad1 (7)
- 2 (6)
- 3 (5)
- 4 (4)
- 5 (3)
- 6 (2)
- Happy7 (1)

| 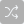 | 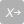 |
| --- | --- |

[sun] On a very bright and completely sunny day how does the sky look?

- Not Cloudy1 (1)
- 2 (2)
- 3 (3)
- 4 (4)
- 5 (5)
- 6 (6)
- Very Cloudy7 (7)

| 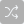 | 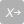 |
| --- | --- |

[lottery] If someone has a winning lottery ticket do they feel good or bad?

- Bad1 (7)
- 2 (6)
- 3 (5)
- 4 (4)
- 5 (3)
- 6 (2)
- Good7 (1)

End of Block

Qualtrics pdf printout of the pre-test survey. The four attention check questions listed were developed *a priori* in order to minimize the possibility of non-human users answering the questions correctly.

**Supplemental eFigure 2:** Full Survey with Consent Form (Qualtrics Printout)

COVID-19 Full Study

Start of Block: consent

UNIVERSITY OF CALIFORNIA LOS ANGELES
STUDY INFORMATION SHEET  

**Lifestyle Decisions** Investigators at UCLA Health and the Anderson School of Management are conducting a research study. You were selected as a possible participant in this study because you are an adult who is a participating member of Mechanical Turk. Your participation in this research study is voluntary.  

**Why is this study being done?** In this study, we want to find out more about the ways that people make decisions. The purpose of this study is to learn how people decide when to go out and what kinds of social contact are important to them. Many of our questions will bring up COVID-19, a new disease caused by a novel coronavirus that has not previously been seen in humans.  

**What will happen if I take part in this research study and how long will it take?** If you volunteer to participate in this study, the researcher will ask you to take a survey about your lifestyle preferences that will take less than 5 minutes.   

**Are there any potential risks or discomforts that I can expect from this study?** There are no anticipated risks or discomforts. Are there any potential benefits if I participate? You will not directly benefit from your participation in the study. The results of the research may aid the understanding of decision making.  

**Will I be paid for participating?** You will receive $.60 for participating.  

**Will information about me and my participation be kept confidential?** The researchers will do their best to make sure that your private information is kept confidential. Information about you will be handled as confidentially as possible, but participating in research may involve a loss of privacy and the potential for a breach in confidentiality. Study data will be physically and electronically secured.  As with any use of electronic means to store data, there is a risk of breach of data security. Your data, including de-identified data may be kept for use in future research.  

**What are my rights if I take part in this study?** You can choose whether or not you want to be in this study, and you may withdraw your consent and discontinue participation at any time. Whatever decision you make, there will be no penalty to you, and no loss of benefits to which you were otherwise entitled. You may refuse to answer any questions that you do not want to answer and still remain in the study.  
**Who can I contact if I have questions about this study?** The research team: If you have any questions, comments or concerns about the research, you can talk to the one of the researchers. Please contact:  Daniel Croymans (310) 206-8000  Kate Christensen (323) 325-1402
 
**UCLA Office of the Human Research Protection Program (OHRPP):** If you have questions about your rights as a research subject, or you have concerns or suggestions and you want to talk to someone other than the researchers, you may contact the UCLA OHRPP by phone: (310) 206-2040; by email:  or by mail: Box 951406, Los Angeles, CA 90095-1406.

- I want to proceed (1)
- I do not want to proceed (2)

End of Block: consent

Start of Block: Intro

Intro In this study we will ask you to imagine yourself within a specific medical scenario.

Please consider the details within your medical scenario when answering the following survey questions.

End of Block: Intro

Start of Block: scenario 1

Scenario 1
Your Medical Scenario:

Imagine that for the past several days you have been experiencing a 
fever, cough, and fatigue.

Your doctor tells you that testing for active COVID-19 viral infection 
is NOT AVAILABLE.

Based on this information, your doctor tells you that you still very likely 
have COVID-19 and that you should self-isolate.

End of Block: scenario 1

Start of Block: scenario 2

Scenario 2
Your Medical Scenario:

Imagine that for the past several days you have been experiencing 
a fever, cough, and fatigue. 

Your doctor then tests you for active COVID-19 viral infection 
and you test POSITIVE.

Based on this information, your doctor tells you that you still very likely 
have COVID-19 and that you should self-isolate.

End of Block: scenario 2

Start of Block: scenario 3

Scenario 3
Your Medical Scenario:

Imagine that for the past several days you have been experiencing 
a fever, cough, and fatigue. 

Your doctor then tests you for active COVID-19 viral infection 
and you test NEGATIVE.

Based on this information, your doctor tells you that you still very likely 
have COVID-19 and that you should self-isolate.

End of Block: scenario 3

Start of Block: activities

Q376 Timing

First Click (1)

Last Click (2)

Page Submit (3)

Click Count (4)

Display This Question:

If Your Medical Scenario 1: Imagine that for the past several days you have been experiencing a  fever... Is Displayed (All scenario information from the prior page is also shown on this page)

Imagine you have the upcoming activities over the following 2 weeks.  How likely would you be to...

Display This Question:

If Your Medical Scenario 2:Imagine that for the past several days you have been experiencing  a fever,... Is Displayed (All scenario information from the prior page is also shown on this page)

Imagine you have the upcoming activities over the following 2 weeks.  How likely would you be to...

Display This Question:

If Your Medical Scenario 3: Imagine that for the past several days you have been experiencing  a fever,... Is Displayed (All scenario information from the prior page is also shown on this page)

Imagine you have these upcoming activities scheduled over the following 2 weeks. How likely would you be to...

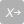

[hair] Go to the barber shop or hair salon to get my haircut.

- Extremely unlikely (1)
- Unlikely (2)
- Somewhat unlikely (3)
- Neither likely nor unlikely (4)
- Somewhat likely (5)
- Likely (6)
- Extremely likely (7)

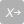

[wedding] Go to my best friend’s wedding at a hotel banquet hall.

- Extremely unlikely (1)
- Unlikely (2)
- Somewhat unlikely (3)
- Neither likely nor unlikely (4)
- Somewhat likely (5)
- Likely (6)
- Extremely likely (7)

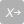

[funeral] Go to a family member’s funeral at a funeral home.

- Extremely unlikely (1)
- Unlikely (2)
- Somewhat unlikely (3)
- Neither likely nor unlikely (4)
- Somewhat likely (5)
- Likely (6)
- Extremely likely (7)

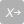

[birthday] Go to a friend or family member’s birthday party at a local park.

- Extremely unlikely (1)
- Unlikely (2)
- Somewhat unlikely (3)
- Neither likely nor unlikely (4)
- Somewhat likely (5)
- Likely (6)
- Extremely likely (7)

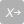

[vote] Vote in-person at my polling place for a local, state, or national election.

- Extremely unlikely (1)
- Unlikely (2)
- Somewhat unlikely (3)
- Neither likely nor unlikely (4)
- Somewhat likely (5)
- Likely (6)
- Extremely likely (7)

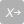

[protest] Attend a political rally or civil protest/ counter-protest.

- Extremely unlikely (1)
- Unlikely (2)
- Somewhat unlikely (3)
- Neither likely nor unlikely (4)
- Somewhat likely (5)
- Likely (6)
- Extremely likely (7)

End of Block: activities

Start of Block: behaviors

Q375 Timing

First Click (1)

Last Click (2)

Page Submit (3)

Click Count (4)

Display This Question:

If Your Medical Scenario 1: Imagine that for the past several days you have been experiencing a  fever... Is Displayed (All scenario information from the first page is also shown on this page

How likely are you to engage in each of these behaviors over the following 2 weeks?

Display This Question:

If Your Medical Scenario 2: Imagine that for the past several days you have been experiencing  a fever,... Is Displayed (All scenario information from the first page is also shown on this page

How likely are you to engage in each of these behaviors over the following 2 weeks?

Display This Question:

If Your Medical Scenario 3: Imagine that for the past several days you have been experiencing  a fever,... Is Displayed (All scenario information from the prior page is also shown on this page

How likely are you to engage in each of these behaviors over the following 2 weeks?

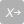

[mask] Wear a face mask anytime I leave my home.

- Extremely unlikely (1)
- Unlikely (2)
- Somewhat unlikely (3)
- Neither likely nor unlikely (4)
- Somewhat likely (5)
- Likely (6)
- Extremely likely (7)

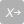

[self-isolate] Self-isolate at home while I have symptoms.

- Extremely unlikely (1)
- Unlikely (2)
- Somewhat unlikely (3)
- Neither likely nor unlikely (4)
- Somewhat likely (5)
- Likely (6)
- Extremely likely (7)

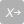

[visit] Visit a friend or family member in person.

- Extremely unlikely (1)
- Unlikely (2)
- Somewhat unlikely (3)
- Neither likely nor unlikely (4)
- Somewhat likely (5)
- Likely (6)
- Extremely likely (7)

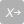

[supplies] Go out to get needed supplies like food and groceries.

- Extremely unlikely (1)
- Unlikely (2)
- Somewhat unlikely (3)
- Neither likely nor unlikely (4)
- Somewhat likely (5)
- Likely (6)
- Extremely likely (7)

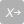

[physicalactivity] Go outside for any type of physical activity (e.g. walk, run, cycling, etc).

- Extremely unlikely (1)
- Unlikely (2)
- Somewhat unlikely (3)
- Neither likely nor unlikely (4)
- Somewhat likely (5)
- Likely (6)
- Extremely likely (7)

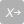

[restaurant] Eat indoors in a restaurant.

- Extremely unlikely (1)
- Unlikely (2)
- Somewhat unlikely (3)
- Neither likely nor unlikely (4)
- Somewhat likely (5)
- Likely (6)
- Extremely likely (7)

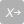

[friendsathome] Invite friends over for dinner inside my home.

- Extremely unlikely (1)
- Unlikely (2)
- Somewhat unlikely (3)
- Neither likely nor unlikely (4)
- Somewhat likely (5)
- Likely (6)
- Extremely likely (7)

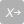

[transportation] Avoid using public transportation.

- Extremely unlikely (1)
- Unlikely (2)
- Somewhat unlikely (3)
- Neither likely nor unlikely (4)
- Somewhat likely (5)
- Likely (6)
- Extremely likely (7)

End of Block: behaviors

Start of Block: Demographics

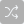

[attention] In your medical scenario, you were diagnosed with COVID-19. Please indicate whether you received testing or not.

- no, test **NOT AVAILABLE** (1)
- yes, tested **POSITIVE** (2)
- yes, tested **NEGATIVE** (3)

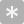

[age] What is your age?

________________________________________________________________

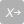

[gender] What best describes your gender?^4^

- Male (1)
- Female (2)
- Prefer to self-describe (3) ________________________________________________
- Prefer not to say (4)

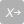

[Hispanic] Are of Spanish, Hispanic, or Latino origin or descent?^5^

- Yes (1)
- No (2)
- Prefer not to say (3)

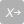

[race] What is your race? Please choose one or more.^5^

- White (1)
- Black or African American (2)
- Asian (3)
- Native Hawaiian or Pacific Islander (4)
- American Indian or Alaskan Native (5)
- Some other race, ethnicity, or origin (6)
- Prefer not to say (7)

| 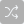 | 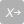 |
| --- | --- |

[politics] In politics, as of today, do you consider yourself a Republican, a Democrat or an independent?^6^

- Republican (1)
- Democrat (2)
- Independent (3)

| Page Break |
| --- |

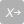

[education] What is the highest grade or level of school that you have completed?^5^

- 8th grade or less (1)
- Some high school, but did not graduate (2)
- High school graduate or GED (3)
- Some college or 2-year degree (4)
- 4-year college degree (5)
- More than 4-year college degree (6)

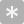

[zip] What is the zip code that you currently reside in?

________________________________________________________________

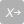

[residence] What type of residence do you live in?^7^

- House/condo/townhouse (1)
- Shelter (2)
- Apartment (3)
- Dormitory (4)
- Assisted living facility (5)
- Skilled nursing center (6)
- No consistent primary residence (7)
- Other (8)

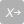

[people] Including yourself, how many people share your kitchen or living space?^7^

- I live by myself (1)
- 2 people (2)
- 3 people (3)
- 4 people (4)
- 5 people (5)
- 6 or more people (6)

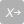

[health] In general, how would you rate your overall health?^5^

- Excellent (1)
- Very good (2)
- Good (3)
- Fair (4)
- Poor (5)

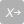

[kids] Is there anyone under the age of 18 living with you?^8^

- Yes (1)
- No (2)

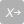

[householdrisk] Is there anyone in your household living with any of the following conditions: age 65 or older, chronic lung disease (e.g. COPD, asthma), heart disease, chronic kidney disease, liver disease, diabetes, or is immunocompromised (e.g. active cancer treatment, immune deficiencies, prolonged corticosteroid use)?^7^

- Yes (1)
- No (2)
- Do not know (3)
- Prefer not to say (4)

| Page Break |
| --- |

Q355 **How well does this statement describe you or your situation?**^9^

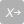

[cfpb1] Because of my money situation, I feel like I will never have the things I want in life.

- Completely (1)
- Very well (2)
- Somewhat (3)
- Very little (4)
- Not at all (5)

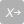

[cfpb2] I am just getting by financially.

- Completely (1)
- Very well (2)
- Somewhat (3)
- Very little (4)
- Not at all (5)

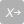

[cfpb3] I am concerned that the money I have or will save won’t last.

- Completely (1)
- Very well (2)
- Somewhat (3)
- Very little (4)
- Not at all (5)

Q360 **How often does this statement apply to you?**

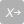

[cfpb4] I have money left over at the end of the month.

- Always (1)
- Often (2)
- Sometimes (3)
- Rarely (4)
- Never (5)

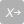

[cfpb5] My finances control my life.

- Always (1)
- Often (2)
- Sometimes (3)
- Rarely (4)
- Never (5)

| Page Break |
| --- |

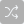

Q405 In your medical scenario, what disease was discussed?

- Hepatitis C (1)
- Herpes (2)
- Syphilis (3)
- Ebola (4)
- HIV/AIDS (5)
- Chlamydia (6)
- COVID-19 (7)
- Tuberculosis (TB) (8)

End of Block: Demographics

Qualtrics printout of the main study survey. Questions regarding age, zip code, and attention check (Q405) were developed *a priori*

**Supplemental eTable 1:** EFA Analysis Results from the 2 previous pilots

| **Table. Exploratory Factor Analysis Results** | | |
| --- | --- | --- |
|  | Factor 1 (Personal) | Factor 2 (Social) |
| friendsathome | 0.73 | 0.42 |
| visit | 0.72 | 0.46 |
| restaurant | 0.71 | 0.35 |
| self_isolate | 0.61 | 0.19 |
| mask | 0.57 | 0.23 |
| physicalactivity | 0.51 | 0.28 |
| supplies | 0.51 | 0.29 |
| birthday | 0.52 | 0.65 |
| hair | 0.57 | 0.63 |
| wedding | 0.39 | 0.83 |
| funeral | 0.26 | 0.78 |
| The two-factor solution was selected as best-fitting based on examination of the Screen plot. 33% of the variance was explained by factor 1, and an additional 26% by factor 2. Cronbach alphas were 0.87 for factor 1, and 0.91 for factor 2, indicating high reliability of the subscores. The alpha for the total score was 0.92.  **Supplemental eFigure 3**: Survey times from 15 college-aged individuals (abridged csv file format)  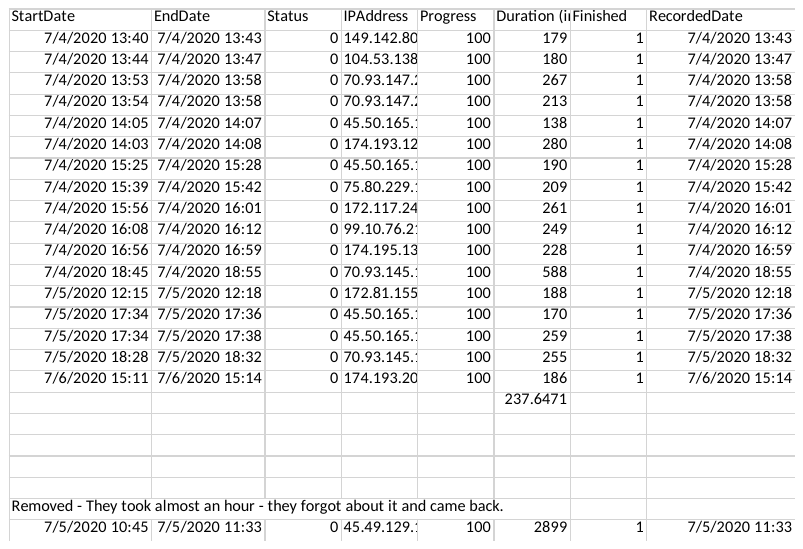  Survey responses from 18 college-educated individuals with time to from beginning to completion listed in “Duration”. One response was removed given that the time to completion was significantly longer than all other responses. The shortest time of 138 seconds was ultimately used by the study team to decide on the minimum 120-second inclusion criteria. | | |

**Supplemental eTable 2:** Total Score, Personal Decisions Subscore, and Social Expectations Subscore Components

| **Personal Decisions Subscore** | **Social Expectations Subscore** | **Individual Items** |
| --- | --- | --- |
| Masking^1^  Self-isolation^2^  Visiting friends^1^  Purchasing supplies^1^ Undertaking physical activity^1^  Eating at a restaurant^1^  Having dinner at home with friends^1^ | Haircut  Attending wedding^1^  Attending Funerals  Birthday party in the park^1^ | Voting  Protesting/Political Rally  Public Transportation^1^ |

Questions regarding haircut, attending a funeral, voting in person, and protesting/political rally were developed *a priori* based on the consensus of the study team. All others behaviors were selected from the CDC website.^1,2^

**Supplemental eTable 3: Post-Hoc Linear Regression Analysis with Individual Construct Scores**

|  | Total Score (R^2^ = 0.36) | | |
| --- | --- | --- | --- |
| Effect | Estimate | SE | P |
| Attitude1 (+1) | -0.12 | 0.05 | **0.009** |
| Attitude2 (+1) | -0.15 | 0.03 | **<0.001** |
| SubjNorm1 (+1) | -0.27 | 0.05 | **<0.001** |
| SubjNorm2 (+1) | -0.04 | 0.02 | 0.147 |
| Control (+1) | -0.05 | 0.02 | **0.022** |

Individual TPB Construct Scores along with mean difference and standard error (SE). Statistically significant differences were denoted with bolded p-values.

**Supplemental eTable 4: Mean Scores and Relative Percent Differences for Total Score, Subscores, and Individual Items**

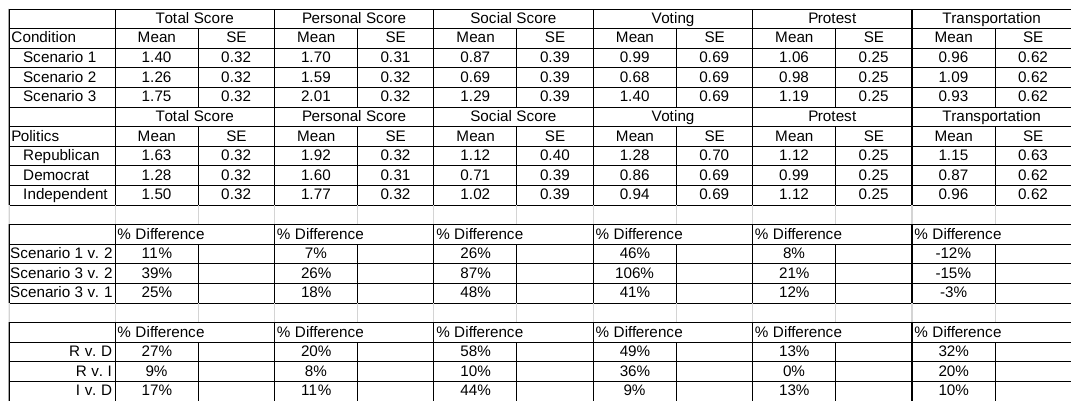

Table above refers to least-square mean scores (mean) and standard error (SE) based on the pre-registered model. “% Difference” refers to the relative difference in risky behavioral intentions as compared to the reference group based on the calculated mean scores. “R” refers to participants who identified as Republican, “D” refers to participants who identified as Democrats, and “I” refers to those who identify as Independents.

**Supplemental eFigure 4: Screenshot of Participant Survey Screen**

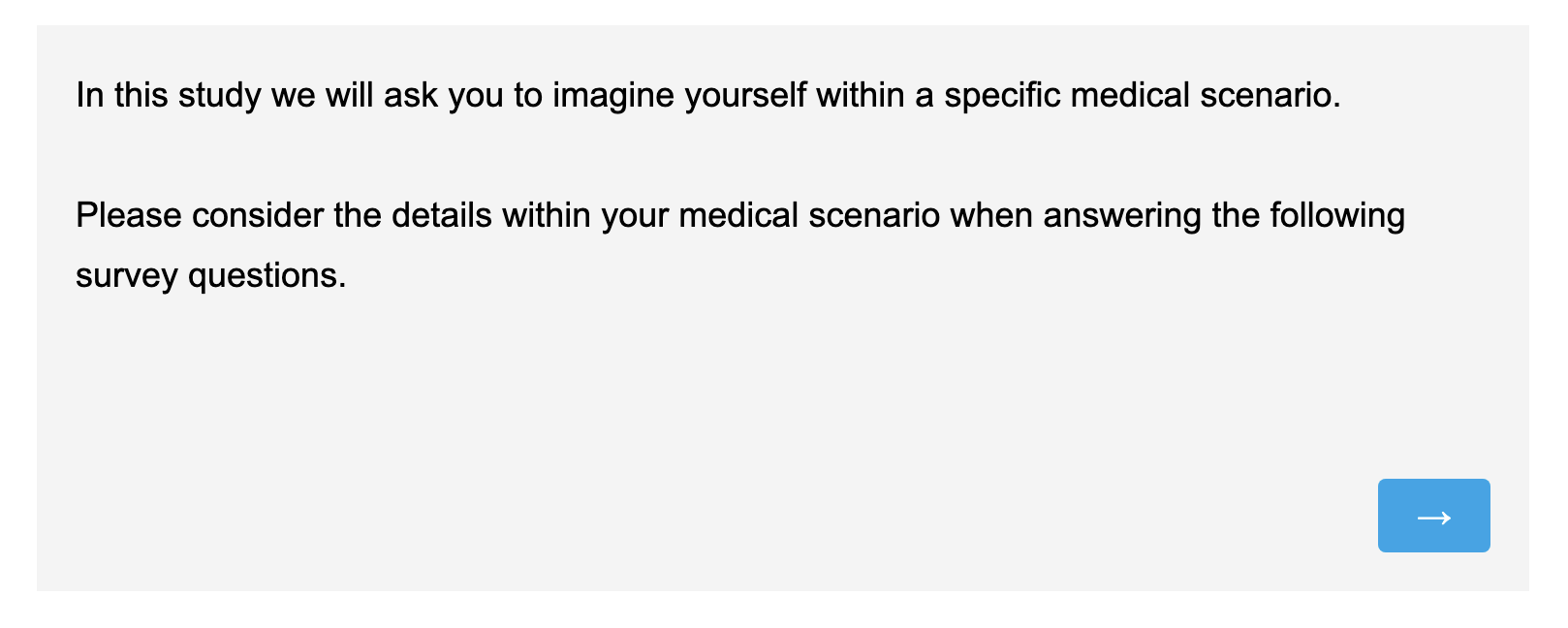

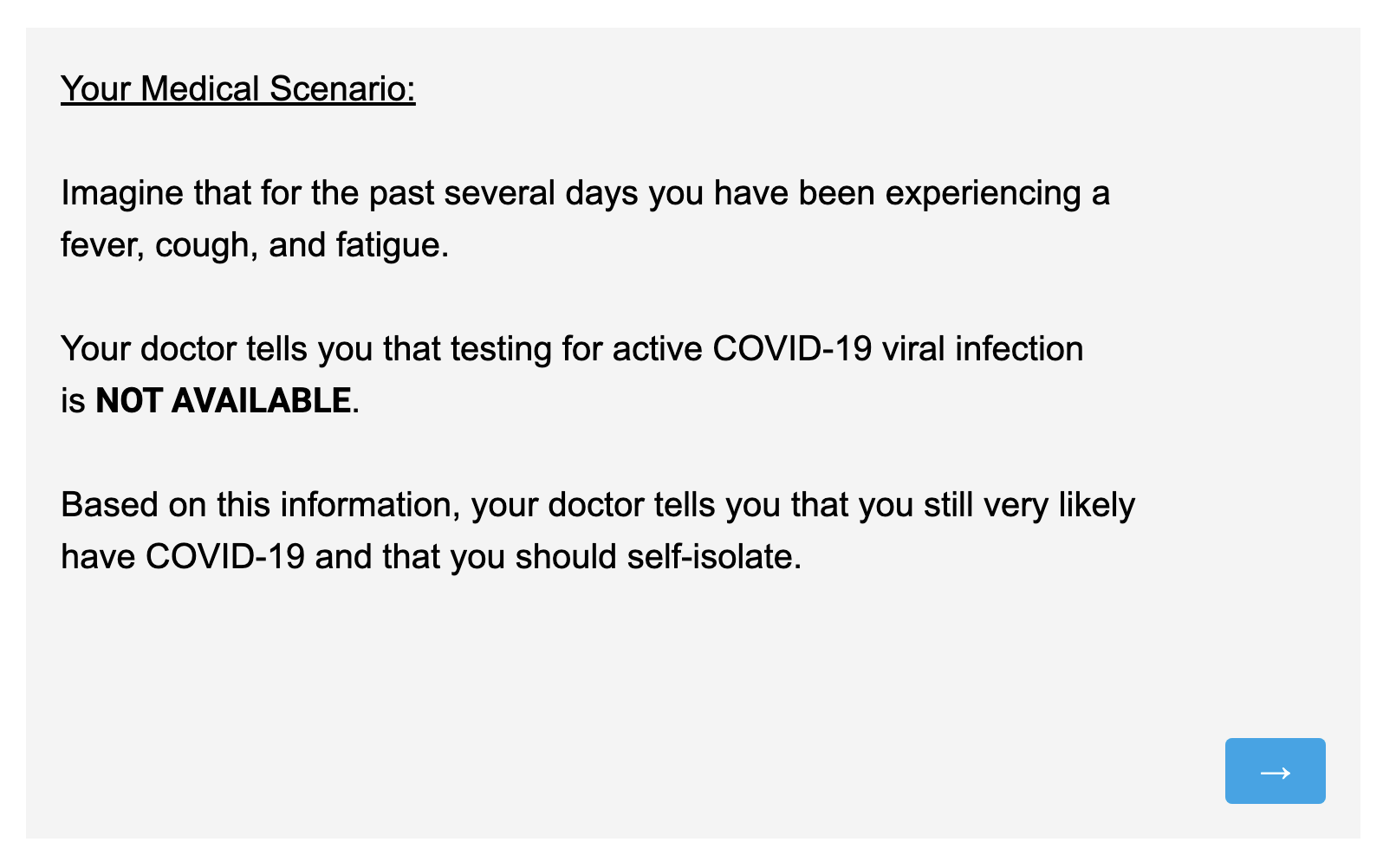

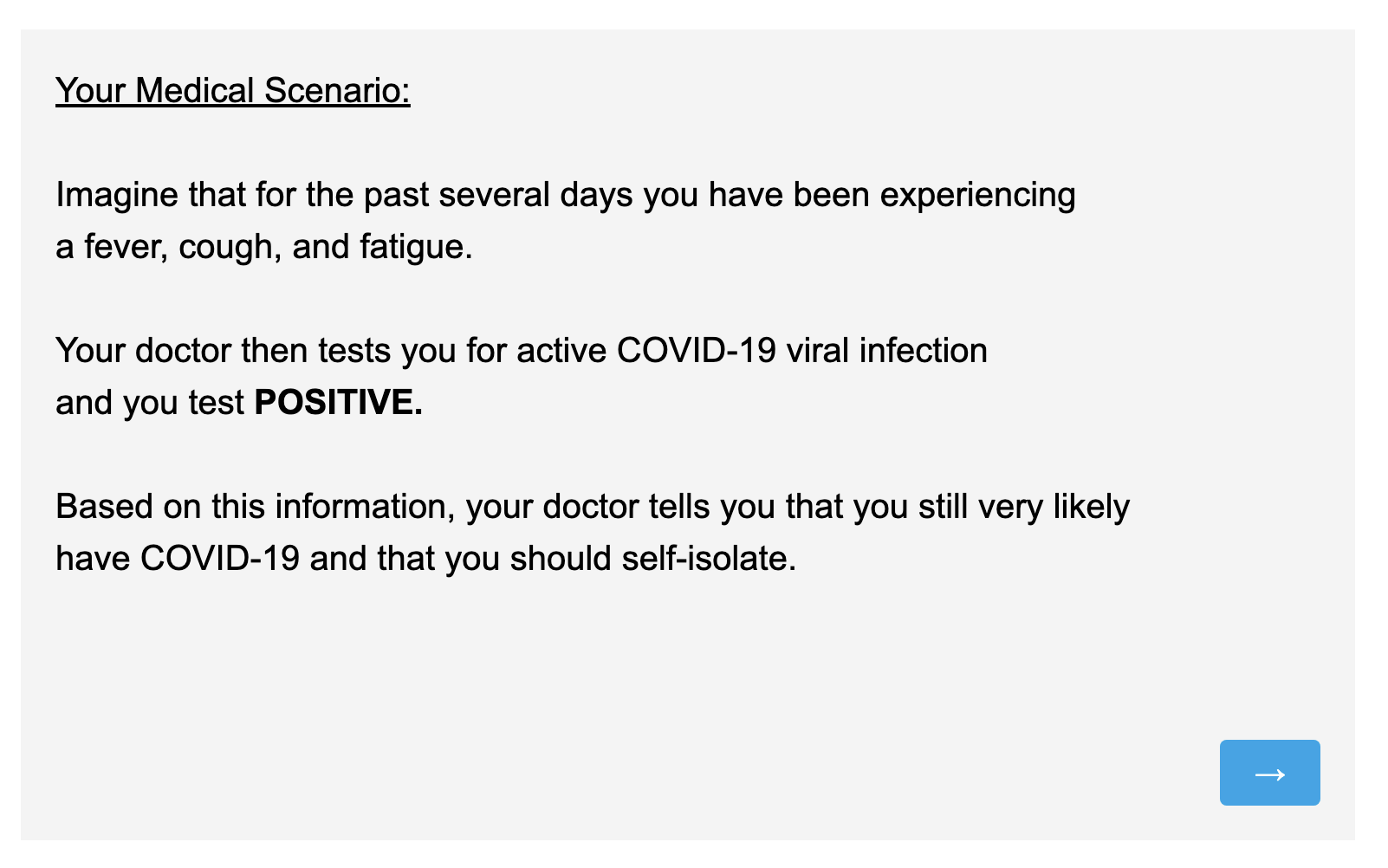

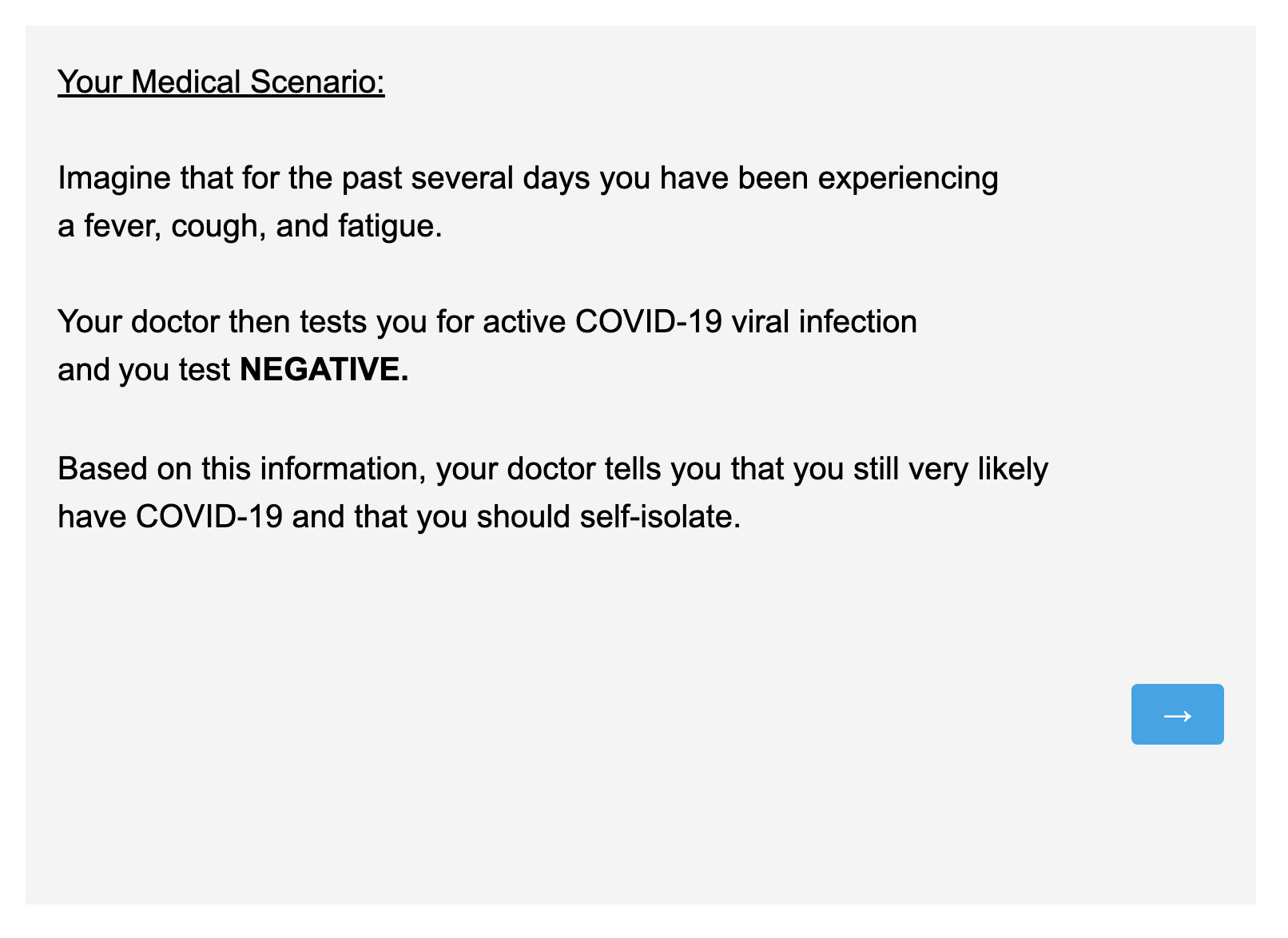

Screenshot images depicting each of the screens that participants saw during the survey. The top image was the introduction screen seen by all participants, while subsequent images were seen by Arm 1, Arm 2, and Arm 3 respectively.
